# Ocrelizumab B cell depletion has no effect on HERV RNA expression in PBMC in MS patients

**DOI:** 10.1101/2023.11.10.23298370

**Authors:** Rachael Tarlinton, Radu Tanasescu, Claire Shannon-Lowe, Bruno Gran

## Abstract

**Background:** Epstein Barr Virus (EBV) infection of B cells is now understood to be one of the triggering events for the development of Multiple Sclerosis (MS), a progressive immune-mediated disease of the central nervous system. EBV infection is also linked to expression of Human endogenous retroviruses (HERVs) of the HERV-W group, a further risk factor for the development of MS (Ocrelizumab is a high-potency disease-modifying treatment (DMT) for MS, which depletes B cells by targeting CD20.

**Objectives:** We studied the effects of ocrelizumab on gene expression in peripheral blood mononuclear cells (PBMC) from paired samples from 20 patients taken prior to and 6 months after beginning Ocrelizumab therapy. We hypothesised that EBV and HERV-W loads would be lower in post-treatment samples.

**Methods:** Samples were collected in Paxgene tubes, subject to RNA extraction and Illumina paired end short read mRNA sequencing with mapping of sequence reads to the human genome using Salmon and Differential Gene expression compared with DeSeq2. Mapping was also performed separately to the HERV-D database of HERV sequences and the EBV reference sequence.

**Results:** Patient samples were more strongly clustered by individual rather than disease type (relapsing/remitting or primary progressive), treatment (pre and post), age, or sex. Fourteen genes, all clearly linked to B cell function were significantly down regulated in the post treatment samples. Interestingly only one pre-treatment sample had detectable EBV RNA and there were no significant differences in HERV expression (of any group) between pre- and post-treatment samples.

**Conclusions:** While EBV and HERV expression are clearly linked to triggering MS pathogenesis, it does not appear that high level expression of these viruses is a part of the ongoing disease process or that changes in virus load are associated with Ocrelizumab treatment.

## Introduction

Multiple sclerosis (MS) is a disabling immune-mediated, inflammatory disease of the central nervous system (CNS) ^1^. Pathological damage is directed against CNS myelin and axons, with clinical manifestations that are characterised by relapses and / or progression of neurological deficits. Most patients present with a relapsing-remitting clinical course (RRMS) characterised by partial recovery between bouts of inflammation, followed by secondary progressive disease (SPMS) in which gradual neurological decline is independent of relapses. A smaller number of patients have progressive disease from the start (primary progressive course, PPMS). In spite of variations in clinical presentation, MS is considered a single disease. It is likely that different pathological processes underlie relapses and progression. Numerous immunotherapies have been approved for the treatment of relapsing-remitting disease and only two for progressive disease. There is general consensus that early treatment of MS is advantageous ^2,3^.

While not solely responsible for MS pathogenesis, viral infections are a known risk factor for the development of the disease and indeed the first established effective therapy for MS was interferon beta, a cytokine with a central role in antiviral immune responses ^4^. In particular, post pubertal infection with Epstein Barr Virus (EBV), normally a common childhood infection, is strongly linked to disease risk and exposure to the virus appears to be required for disease development ^5^. Recent large clinical studies including one of repeated long term blood samples from over 10 million US military personnel have demonstrated this epidemiological link beyond doubt ^6,7^. Over-expression of human endogenous retroviruses (HERVs) of the HERV-W family is also associated with MS disease risk ^8^. These HERVs are copies of ancestral viral infections that have become integrated into the host’s genome and now perform essential host functions (one HERV-W protein is an essential component of the human placenta) ^8^.

There is a rapidly increasing body of evidence linking EBV and HERV (particularly the HERV-W family) proteins with MS pathogenesis. EBV replicates primarily in B cells,^9^ where it establishes latency, associated with the viral protein Epstein Barr nuclear antigen 2 (EBNA2). EBNA2 binds to genetic loci associated with MS risk competing for transcription binding sites with Vitamin D, high levels of which are protective against MS risk and inhibit B cell proliferation ^9^. HERV-W env is expressed specifically in monocytes, T and B lymphocytes and NK cells and is particularly associated with activation of the non-classical monocyte class (CD14^low^CD16^+^) that are upregulated in MS ^10-12^.

EBV replication in B cells triggers HERV-W and HERV-K expression ^13-16^ initiating a cascade of stimulation of inflammation ^17-19^ and cross reactivity with myelin oligodendrocyte protein (MOG) ^20^. HERV-W and EBV expression levels are also associated and EBV and HERV-W loads are correlated in MS patients undergoing therapy ^21^. An additional line of evidence linking HERV-W proteins to MS pathogenesis has shown HERV-W expression in microglia (brain-resident myeloid cells) associates with axons inducing a degenerative phenotype resulting in damage to myelinated axons ^22^. HERV-W has also been shown to inhibit oligodendrocyte precursor cell formation and remyelination, an effect that can be blocked by the anti-HERV monoclonal antibody GNbAC1 ^23,24^.

Both Epstein Barr nuclear antigen 1 (EBNA1) and HERV-W env demonstrate binding to the HLADR2 allele that is the strongest genetic predisposition to MS (DRB1(*) 15:01) ^25-29^ and exhibit cross reactivity with myelin components ^27,30^. EBV specific HLA1 responses are more likely in MS patients and these patients also have EBV-specific memory T cells in their cerebrospinal fluid ^31^. BCR sequencing of the B cell complement combined with screening of the sequenced antibodies against EBV and CNS proteins of MS patient and controls has demonstrated clonal amplification of EBNA1 and GlialCAM (a protein and chloride channel regulator in glial cells important in CNS repair mechanisms) cross reactive B cells in the PMBC and CNS of MS patients ^32^. A similar study demonstrated antibodies cross reactive to both EBV and alpha-crystallin B (CRYAB), a molecular chaperone protein involved in glial responses to injury) enriched in MS patients compared with healthy controls ^33^. Genome wide association studies (GWAS) and transcriptome studies of MS patients have repeatedly indicated antiviral proteins as risk factors in disease occurrence and progression ^5,34,35^. Clinical trials of T cell therapy specifically targeting EBV have even begun ^36^ with promising early results for both clinical improvement and decrease in EBV antibody titre.

It is unclear whether these viruses initiate a triggering event creating an aberrant immune response or B cell type that perpetuates itself in the absence of the viral trigger or whether chronic or high viral loads are part of the underlying pathology. There are a range of studies demonstrating that antibody and T cell responses to EBV are consistently higher in MS patients than controls and that these are elevated during relapsing phases of RRMS ^25,30,37-46^. However, EBV nucleic acid in the blood or shed in saliva is usually not associated with MS ^39,42,47-51^ though it can be detected in CNS/Brain samples ^52^.

EBV establishes life-long latency in a subpopulation of memory B cells and there are strong indications that aberrant latency programming in EBV infected cells, indicated by the presence of the EBNA2 protein may be an important factor in the development of MS ^53^. B cell depletion therapies that broadly target B cell such as cladribine, anti CD-52 antibodies (alemtuzumab) and anti CD-20 antibodies (ocrelizumab, rituxumab) have proven effective in control of clinical disease in MS ^54-57^. In some cases these therapies also result in decreased EBV antibody titre and cellular immune responses ^45,58-61^. Those therapies that specifically target naïve and plasma B cells (atacicept) or boost memory T cells such as infliximab (anti TNF-alpha antibody) or lenercept on the other hand enhance disease ^54,56,62^.

Generation of spontaneous lymphoblastoid cell lines (transformed EBV infected B cells) is more common in MS patients (and in other autoimmune diseases) than in healthy controls ^63,64^ and genetic variation in EBV latency associated proteins ^65^ in MS patients has been demonstrated. Expression of the latency-associated protein, EBNA1, is enhanced in B cells from younger patients ^16^ while “age-associated” B cells (which are expanded in older patients) are also expanded in MS patients and altered based on herpesvirus status ^66^. This B cell subset are T-bet/CXCR3 + memory B cells that skew immune responses to a Th1 (viral and intracellular pathogen) cellular immune response. They are neuroinvasive and are associated with EBV reactivation ^67^. This subset of cells can be induced by an atypical latency programme in EBV infected B cells ^68^ and are currently a key suspect in the cellular triggers of MS.

This study sought to address whether targeting the primary site of viral antigen production for EBV and HERV-W proteins (peripheral B cells) by depletion with the monoclonal antibody Ocrelizumab, which specifically targets the B cell surface protein CD-20 ^2^ reduces viral load and is thereby associated with a reduction in MS pathology and clinical disease.

## Materials and Methods

Ethics approval was granted by the University of Nottingham, Faculty of Medicine and Health Sciences Research and Ethical Committee number: MREC 08/H0408/167.

Twenty patients with RRMS or PPMS were recruited prior to beginning Ocrelizumab therapy (Table 1 and Supplementary Information). Patients were recruited from those undergoing routine therapy for MS through the Nottingham University Hospitals NHS trust, including routine clinical assessment of clinical activity (and usually one MRI brain scan a year). Recruitment criteria aimed to be as even as possible while remaining representative of typical MS patients treated with Ocrelizumab within the timeframe of the study. Patients were between 18 and 65 years old, 75% female and 25% male, 75% with RRMS and 25% with PPMS. Patients had no history of other disease modifying therapies (DMTs) (19 patients) or no DMT during the previous 3 months and no treatment with high-dose steroids for MS relapse within the last 30 days (1 patient). This patient had had one dose of Copaxone (glatiramer acetate) several years previously. Seventy five percent of the patients were recently diagnosed (<1 year). A minimum sample size of 13 (pre and post treatment) was estimated for demonstrating significant differences in transcriptomic studies with an FDR of 0.5 and an expected fold change of 4 ^69^.

Five ml of blood was collected into PaxGene tubes (Qiagen). A follow up blood sample was taken 5 months later, when patients had received their first 2 infusions and were reviewed by the MS Team in preparation for the third infusion at 6 months. Blood samples were stored at -20 °C until RNA extraction. RNA extraction was performed with a Paxgene blood RNA kit (Qiagen) as per manufacturer’s protocol. Illumina NovaSeq RNA sequencing with polyA library preparation (150 base pair, paired end reads) was performed by Novogene UK.

**Table 1:** Summary of Patient Demographics.

| Characteristic | Number of Patients |
| --- | --- |
| **Type of Disease** |  |
| Relapsing Remitting MS | 15 |
| Primary Progressive MS | 5 |
| **Sex** |  |
| F | 15 |
| M | 5 |
| **Age Bracket** |  |
| 20-29 | 5 |
| 30-49 | 3 |
| 40-59 | 9 |
| 50-65 | 3 |
| **Time since initial diagnosis** |  |
| <1 year | 15 |
| <2 years | 1 |
| 7-10 years | 2 |
| Unknown | 2 |
| **Time since onset of symptoms** |  |
| <1 year | 5 |
| <2 years | 3 |
| <3 years | 2 |
| 3-7 years | 3 |
| 8-18 years | 3 |
| Unknown | 4 |
| **Total** | **20** |

The resulting data files were trimmed (quality score 30, min length 150 and adapters removed) with FASTP ^70^. Mapped to the unmasked ensembl version of the human genome (GrCh38) with Salmon ^71^. Additional mapping was performed to the EBV reference genome (NC 007605) and a custom Human endogenous retrovirus databases (HERVd) ^72^. Differential gene expression analysis was performed with the DESeq2 ^73^ pipeline implemented in iDEP ^74^ with the parameters: false discovery rate 0.1, min fold change 2, model: treatment and patient. Hierarchical clustering and heat maps were also generated in iDEP.

## Results

Fourteen genes, of which 13 were clearly linked to B cell function, were downregulated in the post treatment samples; no genes were upregulated. Downregulated genes were: IgG chains (IGHM, 2 variants of IGHD, IGHG2), CD79A and CD79B (part of the B cell receptor complex), CD75 (part of the MHC class II antigen presentation complex), BLK (B lymphocyte tyrosine kinase), MS4A1 (B lymphocyte surface molecule involved in B cell differentiation), VPREB3 (pre assembly of B cell receptor), TCL1A (T cell receptor activator, FCER2 (immunoglobulin E receptor – B cell growth factor) and an unknown transcript (ENSG00000288133).

Hierarchical clustering of sequencing data demonstrated very clear clustering by patient ID (Figure 1). This was a much stronger effect than any other factor in the study (including pre or post treatment, age, sex, or type of disease).

**Figure 1:**
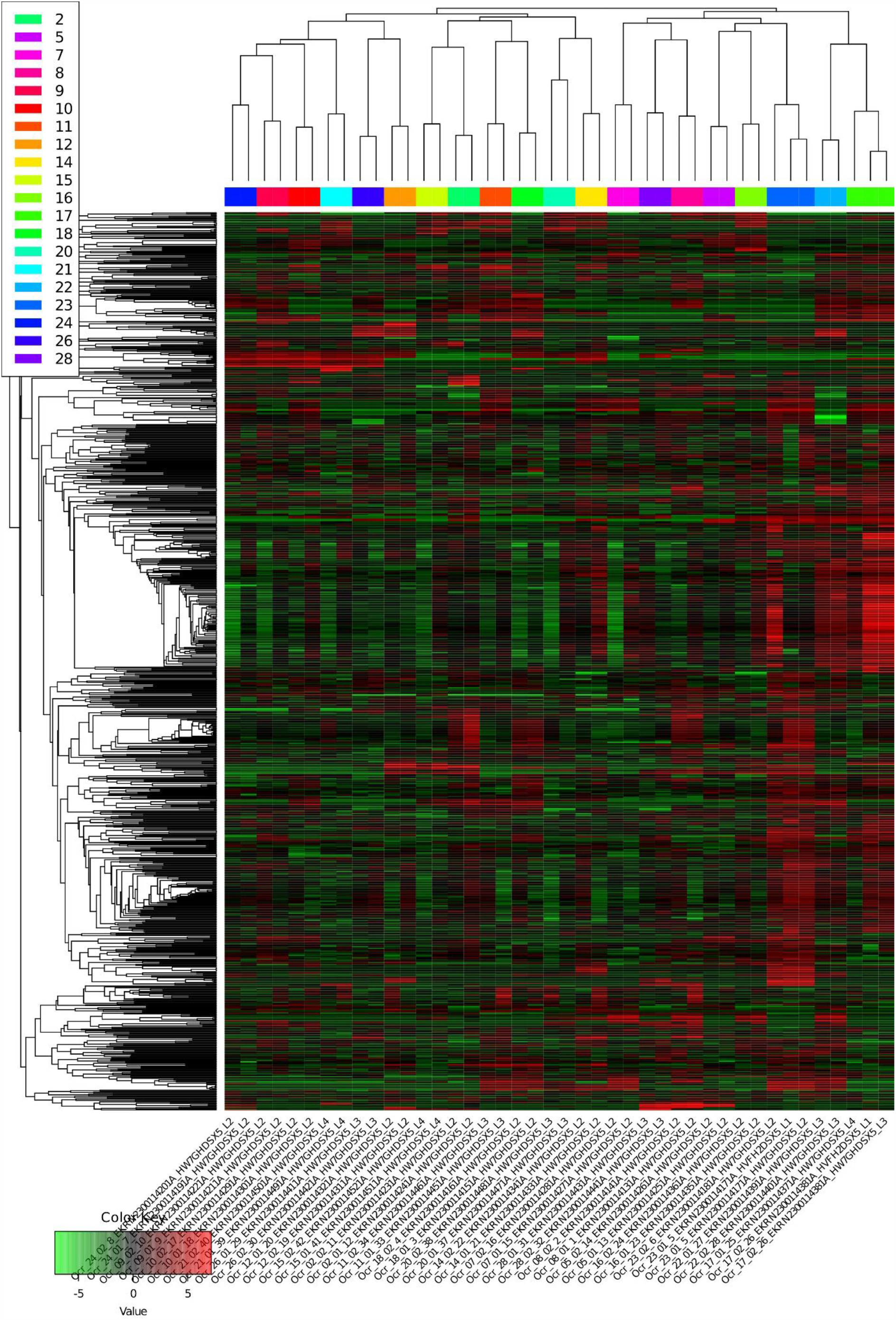
Heat map and Dendogram of hierarchical clustering of patient samples, samples are coloured by patient number (dendrogram) and named by Ocr_patient number_sample number (first or second visit) unique sample ID (two samples were run twice)_unique run ID. Green=lower expression, red=higher expression.

Only one patient had detectable EBV reads (in a pre-treatment sample). Reads for many different HERV groups were detected but there was no differential expression of HERVs in any grouping of patient samples.

## Discussion

MS is a complex disease with the triggering events possibly occurring many years before clinical disease onset and no single antigen target identified ^75^. Between-patient variation is large, necessitating very large epidemiological studies to pin down causal associations like the role of EBV in disease development ^6^. This is reflected in this study where the strongest factor apparent in hierarchical clustering was the individual patient with pre and post treatment samples from the same patient clustering closely (Figure 1). Other factors such as age, sex, type of disease, time since diagnosis or onset of disease had no statistically evident effect on gene expression in this cohort.

With individual patients controlled for in the differential expression model applied to the transcriptomics data the effects of Ocrelizumab B cell depletion were remarkably targeted to a small number of genes (14) with clear B cell associated functions. There have been a small number of other studies using comparable methods and patient cohorts. Fong et al 2023 ^76^ looked at pre and post treatment gene expression in PBMC from 15 Ocrelizumab treated MS patients at 2 weeks and 6 months post therapy compared with 10 healthy controls, 10 untreated MS patients and 9 MS patients treated with Interferon beta, using microarrays. Perhaps unsurprisingly with a more complicated and less controlled cohort and statistical analysis a much larger number of differentially expressed genes (413 decreased and 184 increased) were identified in their study. Similar to our study however the pathways identified were primarily B cell related and 6 of the same genes were identified, namely CD79A, CD22, CD79B, MS4A1 and two IGHD variants. CD22, CD79A and CD79B can be downregulated by EBNA2 and EBNA3 proteins and this affects BCR signalling ^77^. This was suggested to be an additional way through which EBV maintains viral latency and controls the survival of infected B cells ^77^.

Measuring differential expression of HERVs in RNAseq data is not straightforward due to the repetitive nature of transposons, making them not readily distinguishable in some mapping algorithms ^78^. Mapping success is also heavily dependent on the database chosen as the reference sequence. Schwarz et al ^78^ compared the existing algorithms used for this kind of work, TEtranscripts ^79^, SalmonTE ^80^, Telescope ^81^, SQuiRE ^82^and TEtools ^83^ against a known test dataset and found that SalmonTE and Telescope performed reliably. As the original SalmonTE programme is not currently curated, we recreated its functionality using the Salmon mapping algorithm ^71^ and the HERVd ^72^ database of human ERVs, currently the most comprehensive curated database of HERVs. The same approach was taken to EBV mapping. Nali et al ^84^ looked at differential expression of HERVs in 7 MS patients with secondary progressive MS and 3 healthy control PBMCs with Illumina RNAseq and a very similar bio-informatics pipeline but had quite different results to us. HERV-W and 18 additional HERV families were upregulated in MS patients. These differences however probably reflect the different disease stage, smaller number of samples and choice of control in their study (pre-treatment from the same patient in ours vs healthy control in Nali et al^84^).

Similar results to ours indicating a lack of differential expression of HERV transcripts (by RNAseq) in the brains of MS patients and controls were reported by Ekljaer et al 2021 ^85^. As EBV detection in blood is low in normal adults and MS patients ^48^ it is perhaps not surprising that we only detected this in one pre-treatment patient. A lack of differential expression of any HERV family was however unexpected given our original hypothesis that Ocrelizumab depletion of B cells would decrease the opportunity for viral expression in MS patients. This study does however offer support to the body of evidence that suggests that an aberrant cell type or immune response triggered by these viruses, rather than ongoing high viral expression, drives continued pathology in MS. Current evidence points strongly to an aberrant EBV latency programme and resulting in an expanded T-bet/CXCR3 + memory B cell population that is critical in MS pathology ^63,68^.

## Data Availability

RNAseq reads are available at Genbank Bioproject PRJNA1001267 Accession numbers SRR25490470-SRR25490511

## Role of Authors

RTar and BG conceptualised the study, BG and RTan recruited patients, RTar performed the RNA extraction and bio-informatics analysis and drafted the manuscript, All authors reviewed and edited the manuscript.

## References

1 Reich, D. S., Lucchinetti, C. F. & Calabresi, P. A. Multiple Sclerosis. The New England journal of medicine 378, 169–180, doi:10.1056/NEJMra1401483 (2018).

2 Forster, M. et al. Managing Risks with Immune Therapies in Multiple Sclerosis. Drug safety, doi:10.1007/s40264-018-0782-8 (2019).

3 Kuhlmann, T. et al. Multiple sclerosis progression: time for a new mechanism-driven framework. The Lancet. Neurology 22, 78–88, doi:10.1016/s1474-4422(22)00289-7 (2023).

4 Dumitrescu, L., Constantinescu, C. S. & Tanasescu, R. Recent developments in interferon-based therapies for multiple sclerosis. Expert opinion on biological therapy 18, 665–680, doi:10.1080/14712598.2018.1462793 (2018).

5 Afrasiabi, A. et al. Evidence from genome wide association studies implicates reduced control of Epstein-Barr virus infection in multiple sclerosis susceptibility. Genome medicine 11, 26, doi:10.1186/s13073-019-0640-z (2019).

6 Bjornevik, K. et al. Longitudinal analysis reveals high prevalence of Epstein-Barr virus associated with multiple sclerosis. Science 375, 296–301, doi:10.1126/science.abj8222 (2022).

7 Loosen, S. H. et al. Infectious mononucleosis is associated with an increased incidence of multiple sclerosis: Results from a cohort study of 32,116 outpatients in Germany. Frontiers in immunology 13, 937583, doi:10.3389/fimmu.2022.937583 (2022).

8 Morandi, E. et al. The association between human endogenous retroviruses and multiple sclerosis: A systematic review and meta-analysis. PLoS One 12, e0172415, doi:10.1371/journal.pone.0172415 (2017).

9 Ricigliano, V. A. et al. EBNA2 binds to genomic intervals associated with multiple sclerosis and overlaps with vitamin D receptor occupancy. PLoS One 10, e0119605, doi:10.1371/journal.pone.0119605 (2015).

10 Garcia-Montojo, M. et al. Syncytin-1/HERV-W envelope is an early activation marker of leukocytes and is upregulated in multiple sclerosis patients. Eur J Immunol 50, 685–694, doi:10.1002/eji.201948423 (2020).

11 Gjelstrup, M. C. et al. Subsets of activated monocytes and markers of inflammation in incipient and progressed multiple sclerosis. Immunology and cell biology 96, 160–174, doi:10.1111/imcb.1025 (2018).

12 Carstensen, M. et al. Activated monocytes and markers of inflammation in newly diagnosed multiple sclerosis. Immunology and cell biology, doi:10.1111/imcb.12337 (2020).

13 Morandi, E., Tanasescu, R., Tarlinton, R. E., Constantin-Teodosiu, D. & Gran, B. Do Antiretroviral Drugs Protect From Multiple Sclerosis By Inhibiting Expression Of MS-Associated Retrovirus? Frontiers in Immunology. 9, 3092 (2019).

14 Irizar, H. et al. Transcriptomic profile reveals gender-specific molecular mechanisms driving multiple sclerosis progression. PloS one 9, e90482 (2014).

15 Mameli, G. et al. Expression and activation by Epstein Barr virus of human endogenous retroviruses-W in blood cells and astrocytes: inference for multiple sclerosis. PLoS One 7, e44991, doi:10.1371/journal.pone.0044991 (2012).

16 Wieland, L. et al. Epstein-Barr Virus-Induced Genes and Endogenous Retroviruses in Immortalized B Cells from Patients with Multiple Sclerosis. Cells 11, doi:10.3390/cells11223619 (2022).

17 Rolland, A. et al. The envelope protein of a human endogenous retrovirus-W family activates innate immunity through CD14/TLR4 and promotes Th1-like responses. Journal of immunology (Baltimore, Md. : 1950) 176, 7636–7644 (2006).

18 Duperray, A. et al. Inflammatory response of endothelial cells to a human endogenous retrovirus associated with multiple sclerosis is mediated by TLR4. International immunology 27, 545–553, doi:10.1093/intimm/dxv025 (2015).

19 Perron, H. et al. Human endogenous retrovirus protein activates innate immunity and promotes experimental allergic encephalomyelitis in mice. PLoS One 8, e80128, doi:10.1371/journal.pone.0080128 (2013).

20 de Luca, V. et al. Cross-reactivity between myelin oligodendrocyte glycoprotein and human endogenous retrovirus W protein: nanotechnological evidence for the potential trigger of multiple sclerosis. Micron 120, 66–73 (2019).

21 Pérez-Pérez, S. et al. Epstein-Barr Virus Load Correlates with Multiple Sclerosis-Associated Retrovirus Envelope Expression. Biomedicines 10, doi:10.3390/biomedicines10020387 (2022).

22 Kremer, D. et al. pHERV-W envelope protein fuels microglial cell-dependent damage of myelinated axons in multiple sclerosis. Proc Natl Acad Sci U S A 116, 15216–15225, doi:10.1073/pnas.1901283116 (2019).

23 Kremer, D. et al. The neutralizing antibody GNbAC1 abrogates HERV-W envelope protein-mediated oligodendroglial maturation blockade. Multiple sclerosis (Houndmills, Basingstoke, England), doi:10.1177/1352458514560926 (2014).

24 Göttle, P. et al. Rescuing the negative impact of human endogenous retrovirus envelope protein on oligodendroglial differentiation and myelination. Glia 67, 160–170, doi:10.1002/glia.23535 (2019).

25 Hedström, A. K. et al. High Levels of Epstein-Barr Virus Nuclear Antigen-1-Specific Antibodies and Infectious Mononucleosis Act Both Independently and Synergistically to Increase Multiple Sclerosis Risk. Frontiers in neurology 10, 1368, doi:10.3389/fneur.2019.01368 (2019).

26 Ramasamy, R., Joseph, B. & Whittall, T. Potential molecular mimicry between the human endogenous retrovirus W family envelope proteins and myelin proteins in multiple sclerosis. Immunology letters 183, 79–85, doi:10.1016/j.imlet.2017.02.003 (2017).

27 Ramasamy, R., Mohammed, F. & Meier, U. C. HLA DR2b-binding peptides from human endogenous retrovirus envelope, Epstein-Barr virus and brain proteins in the context of molecular mimicry in multiple sclerosis. Immunology letters 217, 15–24, doi:10.1016/j.imlet.2019.10.017 (2020).

28 Menegatti, J., Schub, D., Schäfer, M., Grässer, F. A. & Ruprecht, K. HLA-DRB1*15:01 is a co-receptor for Epstein-Barr virus, linking genetic and environmental risk factors for multiple sclerosis. Eur J Immunol 51, 2348–2350, doi:10.1002/eji.202149179 (2021).

29 do Olival, G. S. et al. Genomic analysis of ERVWE2 locus in patients with multiple sclerosis: absence of genetic association but potential role of human endogenous retrovirus type W elements in molecular mimicry with myelin antigen. Frontiers in microbiology 4, 172, doi:10.3389/fmicb.2013.00172 (2013).

30 Lünemann, J. D. et al. EBNA1-specific T cells from patients with multiple sclerosis cross react with myelin antigens and co-produce IFN-gamma and IL-2. J Exp Med 205, 1763–1773, doi:10.1084/jem.20072397 (2008).

31 Schneider-Hohendorf, T. et al. Broader Epstein-Barr virus-specific T cell receptor repertoire in patients with multiple sclerosis. J Exp Med 219, doi:10.1084/jem.20220650 (2022).

32 Lanz, T. V. et al. Clonally expanded B cells in multiple sclerosis bind EBV EBNA1 and GlialCAM. Nature 603, 321–327, doi:10.1038/s41586-022-04432-7 (2022).

33 Thomas, O. G. et al. Cross-reactive EBNA1 immunity targets alpha-crystallin B and is associated with multiple sclerosis. Sci Adv 9, eadg3032, doi:10.1126/sciadv.adg3032 (2023).

34 Umeton, R. et al. Multiple sclerosis genetic and non-genetic factors interact through the transient transcriptome. Scientific reports 12, 7536, doi:10.1038/s41598-022-11444-w (2022).

35 Locus for severity implicates CNS resilience in progression of multiple sclerosis. Nature, doi:10.1038/s41586-023-06250-x (2023).

36 Smith, C. & Khanna, R. Adoptive T-cell therapy targeting Epstein-Barr virus as a treatment for multiple sclerosis. Clinical & translational immunology 12, e1444, doi:10.1002/cti2.1444 (2023).

37 Banwell, B. et al. Clinical features and viral serologies in children with multiple sclerosis: a multinational observational study. The Lancet. Neurology 6, 773–781, doi:10.1016/s1474-4422(07)70196-5 (2007).

38 Kvistad, S. et al. Antibodies to Epstein-Barr virus and MRI disease activity in multiple sclerosis. Multiple sclerosis (Houndmills, Basingstoke, England) 20, 1833–1840, doi:10.1177/1352458514533843 (2014).

39 Yea, C. et al. Epstein-Barr virus in oral shedding of children with multiple sclerosis. Neurology 81, 1392–1399, doi:10.1212/WNL.0b013e3182a841e4 (2013).

40 Makhani, N. et al. Viral exposures and MS outcome in a prospective cohort of children with acquired demyelination. Multiple sclerosis (Houndmills, Basingstoke, England) 22, 385–388, doi:10.1177/1352458515595876 (2016).

41 Langer-Gould, A. et al. Epstein-Barr virus, cytomegalovirus, and multiple sclerosis susceptibility: A multiethnic study. Neurology 89, 1330–1337, doi:10.1212/wnl.0000000000004412 (2017).

42 Giess, R. M. et al. Epstein-Barr virus antibodies in serum and DNA load in saliva are not associated with radiological or clinical disease activity in patients with early multiple sclerosis. PLoS One 12, e0175279, doi:10.1371/journal.pone.0175279 (2017).

43 Czarnowska, A. et al. Herpesviridae Seropositivity in Patients with Multiple Sclerosis: First Polish Study. European neurology 80, 229–235, doi:10.1159/000496402 (2018).

44 Jakimovski, D. et al. Higher EBV response is associated with more severe gray matter and lesion pathology in relapsing multiple sclerosis patients: A case-controlled magnetization transfer ratio study. Multiple sclerosis (Houndmills, Basingstoke, England), 1352458519828667, doi:10.1177/1352458519828667 (2019).

45 Persson Berg, L. et al. Serum IgG levels to Epstein-Barr and measles viruses in patients with multiple sclerosis during natalizumab and interferon beta treatment. BMJ Neurol Open 4, e000271, doi:10.1136/bmjno-2022-000271 (2022).

46 Comabella, M. et al. Increased cytomegalovirus immune responses at disease onset are protective in the long-term prognosis of patients with multiple sclerosis. Journal of neurology, neurosurgery, and psychiatry 94, 173–180, doi:10.1136/jnnp-2022-330205 (2023).

47 Lindsey, J. W., Hatfield, L. M., Crawford, M. P. & Patel, S. Quantitative PCR for Epstein-Barr virus DNA and RNA in multiple sclerosis. Multiple sclerosis (Houndmills, Basingstoke, England) 15, 153–158, doi:10.1177/1352458508097920 (2009).

48 Cocuzza, C. E. et al. Quantitative detection of epstein-barr virus DNA in cerebrospinal fluid and blood samples of patients with relapsing-remitting multiple sclerosis. PLoS One 9, e94497, doi:10.1371/journal.pone.0094497 (2014).

49 Mostafa, A. et al. Multiple sclerosis-associated retrovirus, Epstein-Barr virus, and vitamin D status in patients with relapsing remitting multiple sclerosis. Journal of medical virology 89, 1309–1313, doi:10.1002/jmv.24774 (2017).

50 Holden, D. W. et al. Epstein Barr virus shedding in multiple sclerosis: Similar frequencies of EBV in saliva across separate patient cohorts. Mult Scler Relat Disord 25, 197–199, doi:10.1016/j.msard.2018.07.041 (2018).

51 Pereira, J. G. et al. Higher frequency of Human herpesvirus-6 (HHV-6) viral DNA simultaneously with low frequency of Epstein-Barr virus (EBV) viral DNA in a cohort of multiple sclerosis patients from Rio de Janeiro, Brazil. Mult Scler Relat Disord 76, 104747, doi:10.1016/j.msard.2023.104747 (2023).

52 Hassani, A., Corboy, J. R., Al-Salam, S. & Khan, G. Epstein-Barr virus is present in the brain of most cases of multiple sclerosis and may engage more than just B cells. PLoS One 13, e0192109, doi:10.1371/journal.pone.0192109 (2018).

53 Keane, J. T. et al. The interaction of Epstein-Barr virus encoded transcription factor EBNA2 with multiple sclerosis risk loci is dependent on the risk genotype. EBioMedicine 71, 103572, doi:10.1016/j.ebiom.2021.103572 (2021).

54 Furman, M. J. et al. B cell targeted therapies in inflammatory autoimmune disease of the central nervous system. Frontiers in immunology 14, 1129906, doi:10.3389/fimmu.2023.1129906 (2023).

55 Dyer, Z., Tscharke, D., Sutton, I. & Massey, J. From bedside to bench: how existing therapies inform the relationship between Epstein-Barr virus and multiple sclerosis. Clinical & translational immunology 12, e1437, doi:10.1002/cti2.1437 (2023).

56 Gensicke, H. et al. Monoclonal antibodies and recombinant immunoglobulins for the treatment of multiple sclerosis. CNS Drugs 26, 11–37, doi:10.2165/11596920-000000000-00000 (2012).

57 Ho, S. et al. Ocrelizumab Treatment Modulates B-Cell Regulating Factors in Multiple Sclerosis. Neurology(R) neuroimmunology & neuroinflammation 10, doi:10.1212/nxi.0000000000200083 (2023).

58 Pham, H. P. T., Gupta, R. & Lindsey, J. W. The cellular immune response against Epstein-Barr virus decreases during ocrelizumab treatment. Mult Scler Relat Disord 56, 103282, doi:10.1016/j.msard.2021.103282 (2021).

59 Pham, H. P. T., Saroukhani, S. & Lindsey, J. W. The concentrations of antibodies to Epstein-Barr virus decrease during ocrelizumab treatment. Mult Scler Relat Disord 70, 104497, doi:10.1016/j.msard.2023.104497 (2023).

60 Domínguez-Mozo, M. I. et al. Epstein-Barr Virus and multiple sclerosis in a Spanish cohort: A two-years longitudinal study. Frontiers in immunology 13, 991662, doi:10.3389/fimmu.2022.991662 (2022).

61 Zivadinov, R. et al. Effect of ocrelizumab on leptomeningeal inflammation and humoral response to Epstein-Barr virus in multiple sclerosis. A pilot study. Mult Scler Relat Disord 67, 104094, doi:10.1016/j.msard.2022.104094 (2022).

62 Kappos, L. et al. Atacicept in multiple sclerosis (ATAMS): a randomised, placebo-controlled, double-blind, phase 2 trial. The Lancet. Neurology 13, 353–363, doi:10.1016/s1474-4422(14)70028-6 (2014).

63 Soldan, S. et al. Unstable EBV latency drives inflammation in multiple sclerosis patient derived spontaneous B cells. Res Sq, doi:10.21203/rs.3.rs-2398872/v1 (2023).

64 Fraser, K. B., Haire, M., Millar, J. H. & McCrea, S. Increased tendency to spontaneous in-vitro lymphocyte transformation in clinically active multiple sclerosis. Lancet 2, 175–176 (1979).

65 Varvatsi, D., Richter, J., Tryfonos, C., Pantzaris, M. & Christodoulou, C. Association of Epstein-Barr virus latently expressed genes with multiple sclerosis. Mult Scler Relat Disord 52, 103008, doi:10.1016/j.msard.2021.103008 (2021).

66 Mouat, I. C. et al. Gammaherpesvirus infection drives age-associated B cells toward pathogenicity in EAE and MS. Sci Adv 8, eade6844, doi:10.1126/sciadv.ade6844 (2022).

67 Leffler, J. et al. Circulating Memory B Cells in Early Multiple Sclerosis Exhibit Increased IgA(+) Cells, Globally Decreased BAFF-R Expression and an EBV-Related IgM(+) Cell Signature. Frontiers in immunology 13, 812317, doi:10.3389/fimmu.2022.812317 (2022).

68 SoRelle, E. D., Reinoso-Vizcaino, N. M., Horn, G. Q. & Luftig, M. A. Epstein-Barr virus perpetuates B cell germinal center dynamics and generation of autoimmune-associated phenotypes in vitro. Frontiers in immunology 13, 1001145, doi:10.3389/fimmu.2022.1001145 (2022).

69 RnaSeqSampleSize, <https://cqs-vumc.shinyapps.io/rnaseqsamplesizeweb/> (

70 Chen, S., Zhou, Y., Chen, Y. & Gu, J. fastp: an ultra-fast all-in-one FASTQ preprocessor. Bioinformatics 34, i884–i890, doi:10.1093/bioinformatics/bty560 (2018).

71 Patro, R., Duggal, G., Love, M. I., Irizarry, R. A. & Kingsford, C. Salmon provides fast and bias-aware quantification of transcript expression. Nature methods 14, 417–419, doi:10.1038/nmeth.4197 (2017).

72 Paces, J., Pavlícek, A. & Paces, V. HERVd: database of human endogenous retroviruses. Nucleic acids research 30, 205–206, doi:10.1093/nar/30.1.205 (2002).

73 Love, M. I., Huber, W. & Anders, S. Moderated estimation of fold change and dispersion for RNA-seq data with DESeq2. Genome Biol 15, 550, doi:10.1186/s13059-014-0550-8 (2014).

74 Ge, S. X., Son, E. W. & Yao, R. iDEP: an integrated web application for differential expression and pathway analysis of RNA-Seq data. BMC bioinformatics 19, 534, doi:10.1186/s12859-018-2486-6 (2018).

75 Gåsland, H. et al. Antibodies to expanded virus antigen panels show elevated diagnostic sensitivities in multiple sclerosis and optic neuritis. Immunology letters 254, 54–64, doi:10.1016/j.imlet.2023.02.003 (2023).

76 Fong, C. C., Spencer, J., Howlett-Prieto, Q., Feng, X. & Reder, A. T. Adaptive and innate immune responses in multiple sclerosis with anti-CD20 therapy: Gene expression and protein profiles. Frontiers in neurology 14, 1158487, doi:10.3389/fneur.2023.1158487 (2023).

77 Khasnis, S. et al. Regulation of B cell receptor signalling by Epstein-Barr virus nuclear antigens. The Biochemical journal 479, 2395–2417, doi:10.1042/bcj20220417 (2022).

78 Schwarz, R., Koch, P., Wilbrandt, J. & Hoffmann, S. Locus-specific expression analysis of transposable elements. Brief Bioinform 23, doi:10.1093/bib/bbab417 (2022).

79 Jin, Y., Tam, O. H., Paniagua, E. & Hammell, M. TEtranscripts: a package for including transposable elements in differential expression analysis of RNA-seq datasets. Bioinformatics 31, 3593–3599, doi:10.1093/bioinformatics/btv422 (2015).

80 Jeong, H. H., Yalamanchili, H. K., Guo, C., Shulman, J. M. & Liu, Z. An ultra-fast and scalable quantification pipeline for transposable elements from next generation sequencing data. Pac Symp Biocomput 23, 168–179 (2018).

81 Bendall, M. L. et al. Telescope: Characterization of the retrotranscriptome by accurate estimation of transposable element expression. PLoS Comput Biol 15, e1006453, doi:10.1371/journal.pcbi.1006453 (2019).

82 Yang, W. R., Ardeljan, D., Pacyna, C. N., Payer, L. M. & Burns, K. H. SQuIRE reveals locus-specific regulation of interspersed repeat expression. Nucleic acids research 47, e27, doi:10.1093/nar/gky1301 (2019).

83 Lerat, E., Fablet, M., Modolo, L., Lopez-Maestre, H. & Vieira, C. TEtools facilitates big data expression analysis of transposable elements and reveals an antagonism between their activity and that of piRNA genes. Nucleic acids research 45, e17, doi:10.1093/nar/gkw953 (2017).

84 Nali, L. H. et al. Human endogenous retrovirus and multiple sclerosis: A review and transcriptome findings. Mult Scler Relat Disord 57, 103383, doi:10.1016/j.msard.2021.103383 (2022).

85 Elkjaer, M. L. et al. Unbiased examination of genome-wide human endogenous retrovirus transcripts in MS brain lesions. Multiple sclerosis (Houndmills, Basingstoke, England) 27, 1829–1837, doi:10.1177/1352458520987269 (2021).

